# Metabologenomics identified fecal biomarkers for bowel movement regulation by *Bifidobacterium longum* capsules: an RCT

**DOI:** 10.1101/2020.03.23.20041400

**Authors:** Y. Nakamura, S. Suzuki, S. Murakami, K. Higashi, N. Watarai, Y. Nishimoto, J. Umetsu, C. Ishii, Y. Ito, Y. Mori, M. Kohno, T. Yamada, S. Fukuda

## Abstract

**Background:** *Bifidobacterium longum* supplementation can be used to regulate bowel movement; however, individuals vary in the response to *B. longum* treatment. One putative factor is the gut microbiota; recent studies have reported that the gut microbiota mediates diet or drug effects. Here, we investigated intestinal features related to *B. longum* effectiveness in increasing bowel movement frequency.

**Results:** A randomized, double-blind controlled crossover trial was conducted with twenty Japanese subjects selected from 50 participants. The subjects received a two-week dietary intervention consisting of *B. longum* in acid-resistant seamless capsules or similarly encapsulated starch powder. Bowel movement frequency was recorded daily, and time-series fecal collection was conducted for metabologenomic analyses. There were differences among subjects in *B. longum* intake-induced bowel movement frequency. The responders were predictable by machine learning based on the metabologenomic features of the fecal samples collected before *B. longum* intake. Between responders and non-responders, the abundances of nine bacterial genera and of three compounds were significantly different.

**Conclusions:** Thus, the gut microbiome and metabolome composition have a strong impact on *B. longum* supplementation effectiveness in increasing bowel movement frequency, and gut metabologenomics enables *B. longum supplementation effect prediction before intake. These findings have implications for th*e development of personalized probiotic treatments.

**Trial registration:** UMIN-CTR, UMIN000018924. Registered 07 September 2015, https://upload.umin.ac.jp/cgi-open-bin/ctr_e/ctr_view.cgi?recptno=R000021894

## Main text

### Background

Constipation is a globally common gastrointestinal disorder of people of all ages. Constipation has been reported in 5.4% to 26.2% of the general population [1–3] and to predispose individuals to a wide range of diseases, including irritable bowel syndrome (IBS), Parkinson’s disease and kidney disease [4–6]. Several diets are used to relieve constipation. For example, dietary fiber and magnesium have been reported to increase bowel movement frequency [3,7–9]. In addition, in recent years, fermented products such as yoghurt have attracted attention because they are useful in relieving constipation. *Bifidobacterium* is a representative microbial genus of probiotics that is frequently used to regulate bowel movements [10]. Multiple randomized controlled trials have shown that bifidobacteria intake increases bowel movement frequency [11–14] or shortens gut transit time [15,16]. However, the response to bifidobacteria supplementation varies from subject to subject. In a study of patients with IBS, a gastrointestinal disorder that causes constipation, 47% of patients exhibited improved symptoms under *Bifidobacterium* supplementation [11].

One factor that could account for these observations is the composition of the gut microbiota. Indeed, individual differences in gut microbiota composition are known to affect probiotic-mediated stimulation [17]. Recent studies based on high-throughput sequencing of the human microbiome have elucidated how the host response to diet is affected by intestinal microbes [18–23]. For example, gut microbial dysbiosis induced by excessive consumption of artificial sweeteners impairs the host’s glucose tolerance metabolism [18], and excessive dietary emulsifiers promote colitis and metabolic syndrome via the gut microbiome [19]. In addition, a study has reported that the gut microbiome mediates the effect of drugs, with variation among individuals in the strength of this effect [24]. Anticancer drugs are one of these drugs. The effect of an immune checkpoint inhibitor targeting PD-1/PD-L1 has been found to be related to the relative abundances of *Bifidobacterium longum*, *Collinsella aerofaciens* and *Enterococcus faecium* or *Akkermansia muciniphila* [25,26], and the alpha diversity and relative abundance of bacteria of the *Ruminococcaceae* family in responders have been found to be higher than those in nonresponders [27]. In addition, the inhibitory effect of the immune checkpoint on CTLA-4 has been shown to be related to the abundance of *Bacteroides* [28].

These individual differences can be predicted with some accuracy from omics data for the gut microbiome [20,21]. In inflammatory bowel disease (IBD), which is one of the most well-known gastrointestinal disorders, the response to anti-integrin biologic therapy is predictable from gut microbial function [29]. With prediction of drug or diet effects from the state of the intestinal environment, it might be possible to prescribe according to each case. In other words, the companion diagnosis based on the individual intestinal environment could enable personalized health care and medical cost reduction.

In this study, we conducted a randomized double-blind controlled crossover trial with healthy subjects to determine the effect of *B. longum* BB536 supplementation on bowel movement frequency (Figure 1A). In addition, we conducted 16S rRNA metagenomic and metabolomic analyses of fecal samples collected during the trial to evaluate the influence of *B. longum* BB536 supplementation on the intestinal environment. Furthermore, the bowel movement frequency-increasing effect of *B. longum* BB536 intake for each individual was estimated using a Bayesian statistical model.

**Figure 1.**
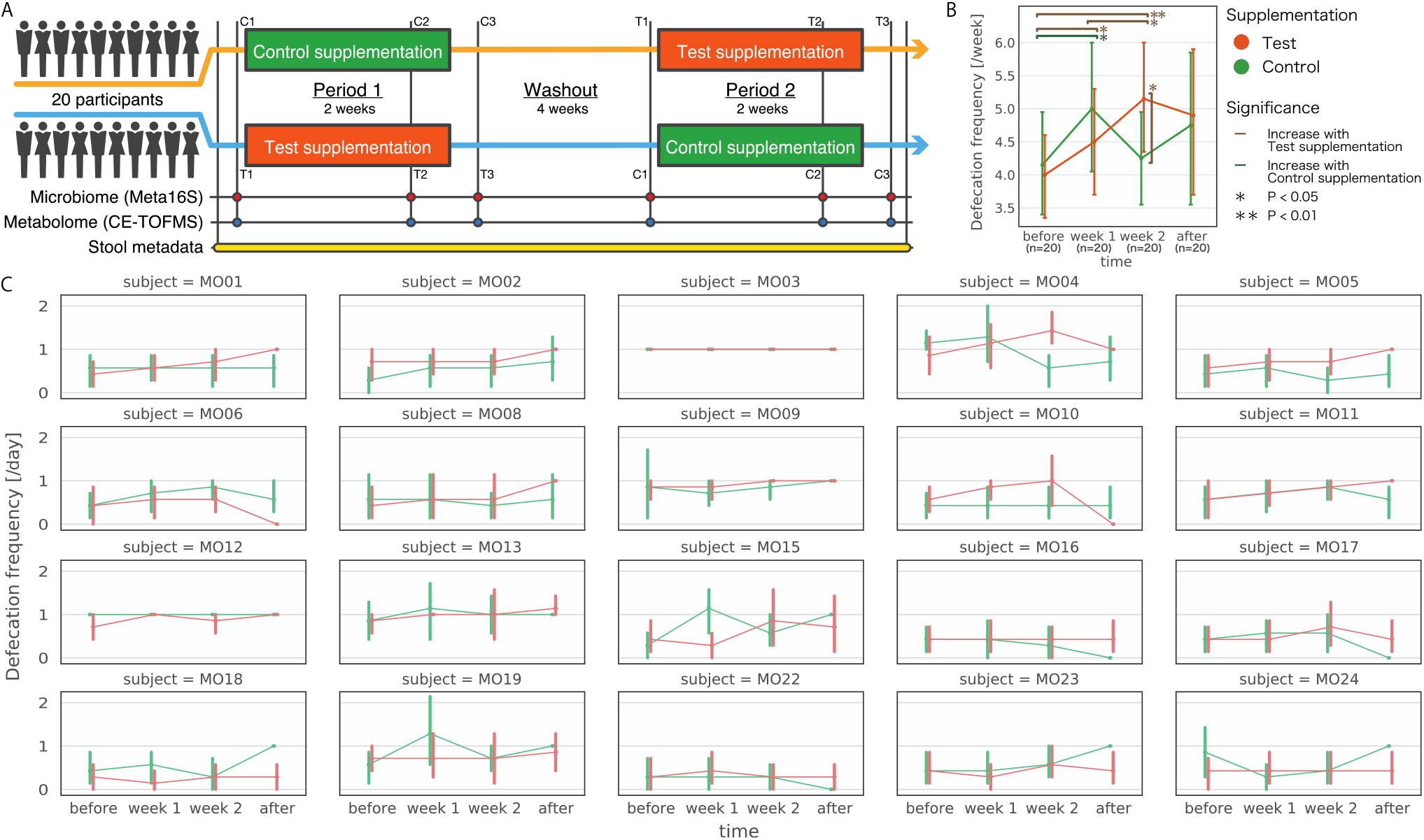
Schematic of the randomized double-blind controlled trial to investigate the increase in bowel movement frequency induced by the intake of *Bifidobacterium longum* BB536 encapsulated in an acid-resistant seamless capsule. A: The illustration indicates a study design of a randomized double-blind controlled trial to investigate the increase in bowel movement frequency due to the intake of *B. longum* BB536 encapsulated in an acid-resistant seamless capsule. B, C: The line graphs indicate the bowel movement frequency per week (B) and the bowel movement frequency per day for each subject (C) throughout the trial period. A Wilcoxon signed rank test (two-sided) was carried out for comparisons between periods (before (n=20); week 1 (n=20); week 2 (n=20); after (n=20)). The points represent the mean bowel movement frequency per week (or day), and the error bars represent the corresponding 95% confidence intervals.

## Results

### The effect of *Bifidobacterium longum* BB536 intake on bowel movement frequency varies among individuals

We confirmed that the bowel movement frequency increased during the test intervention period by the Wilcoxon signed rank test (Figure 1B, Table S1). The bowel movement frequency in the first and second weeks of the test intervention period (4.5 ± 1.9 [/week] and 5.2 ± 2.0 [/week], respectively) increased relative to that in the period before intervention (4.0 ± 1.5 [/week]; p = 0.0323 and p = 0.0012, respectively). In the second week of the test intervention period, the bowel movement frequency increased relative to that in the second week of the control intervention period (4.3 ± 1.7 [/week], p = 0.0335). On the other hand, the bowel movement frequency in the first week of control intervention (5.0 ± 2.2 [/week]) increased relative to that in the period before intervention (4.2 ± 1.8 [/week], p = 0.0335).

In this study, individual variation had observed in the increasing effect of *Bifidobacterium longum* supplementation on bowel movement frequency. Therefore, we analyzed the bowel movement frequency of each individual (Figure 1C). We found that the dynamics of bowel movement frequency over the trial period varied greatly among individuals. For example, in MO04, MO10 and MO11, bowel movement frequency tended to increase over the test intervention period, whereas in MO16, MO18, MO19, MO22, MO23 and MO24, there was little change. Similarly, variation was observed in the control intervention, with some individuals exhibiting increases in bowel movement frequency due to the placebo effect and others not.

### The intestinal environment of each individual was robust for probiotic diet intervention

To determine how the microbiome composition of the subjects changed over the course of the experiment, the 16S rRNA gene region was amplified from the DNA extracted from the fecal samples and sequenced by high-throughput sequencing. Based on these sequencing data, the relative abundance, alpha diversity and beta diversity at the bacterial genus and species level were calculated. Furthermore, metabolites were extracted from the fecal samples and analyzed by CE-TOFMS. Alpha diversity representing the intra-sample complexity of the microbiome composition and beta diversity representing the inter-sample diversity were calculated from the metabolome data. No overall trends were observed in intestinal microbiome and metabolome composition in either of the two intervention groups (Figure 2A, C). The results of multidimensional scaling (MDS) analysis using beta diversity to analyze similarity between all of the samples showed that the microbiome and metabolome composition of the same subject were similar over the course of the experiment (Figure 2B, D, Figure S1A, B). This result suggests that the effects of individual differences in the intestinal microbiome and metabolome composition were larger than the effect of the intervention.

**Figure 2.**
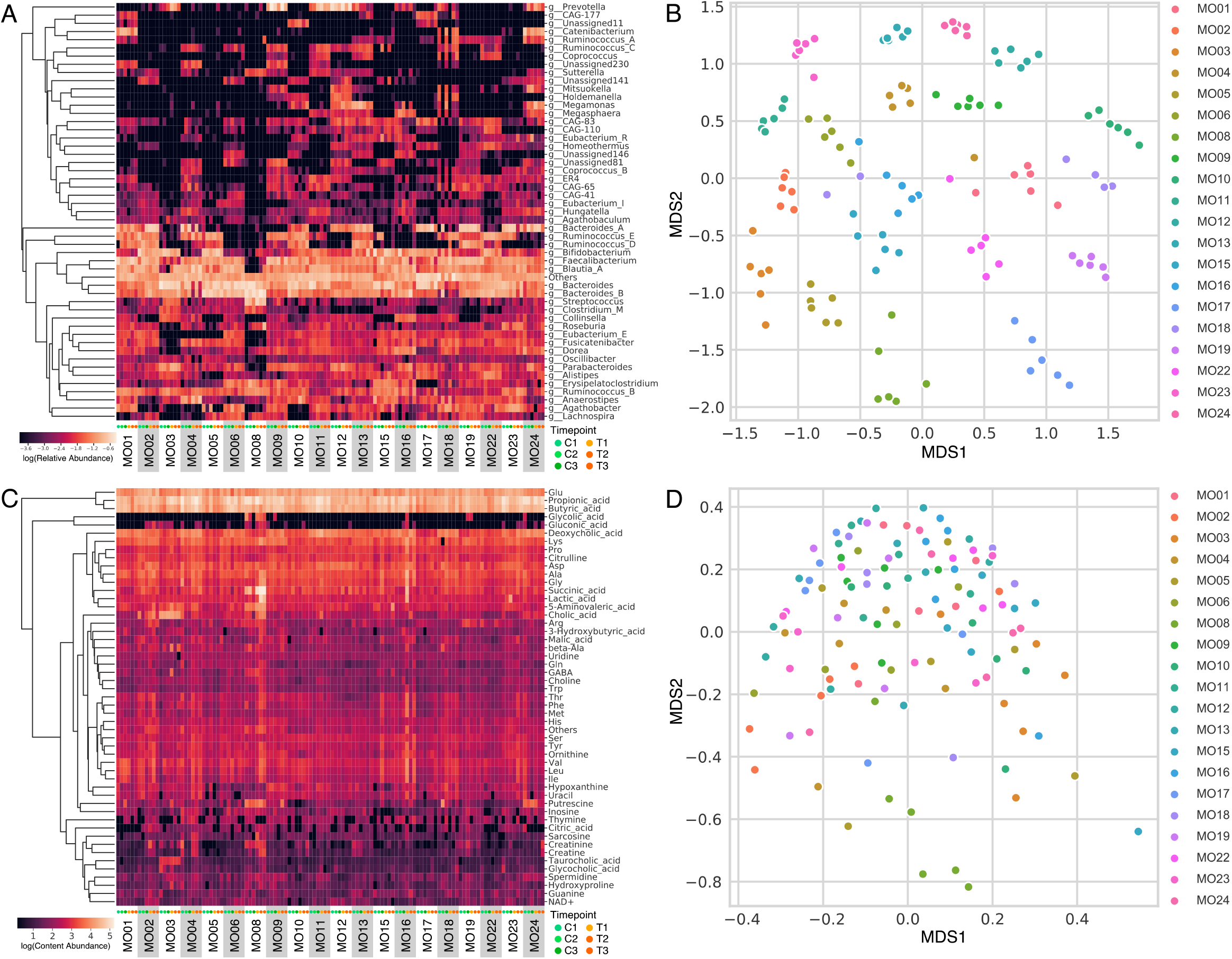
Overall view of the metabologenomic data. A: The heatmap represents the relative abundance of the top 50 genera. B: The scatter plot shows the results of MDS using beta diversity (Spearman correlation coefficient distance) calculated from the microbiome composition. C: The heatmap represents the abundance of the top 50 metabolites in the fecal content. D: A scatter plot showing the results of MDS using beta diversity (Spearman correlation coefficient distance) calculated from the metabolite composition. The same color was used for each subject.

Subsequently, the Brunner-Munzel test (two-sided test) was carried out to compare the changes in gut microbes and metabolites among the test intervention group (T2, T3), the control intervention group (C2, C3), and the nonintervention group (T1, C1). The results showed that some of the bacterial genera were different in the test intervention group from those in the other groups (p value < 0.05, not corrected). However, when false discovery rate (FDR) correction was applied, no changes were detected (Table S2), and no significant change was detected in alpha diversity (Table S3, Figure S2). The relative abundance of *B. longum* tended to increase during the test intervention period and varied among individuals (Figure S3). Overall, our results demonstrate that the influences of *B. longum* BB536 intake on the intestinal microbiome and metabolome were small relative to the influence of individual differences in intestinal environment and that the intestinal environment of each individual was robust for probiotic diet intervention.

### Determining bowel responder by Bayesian Weibull regression

The extent of increase in bowel movement frequency due to *Bifidobacterium longum* supplementation or placebo effect varied among subjects (Figure 1C). To test whether the microbiome or metabolome features of an individual’s intestinal environment varied with the magnitude of response, we first quantified the magnitude of the treatment effect in each individual using a Bayesian statistical model. The models were formulated to estimate the sizes of placebo effect and the test supplement effect precisely. In the models, the bowel movement interval of each individual was used based on the fact that bowel movement tends to occur with the lapse of time. The bowel movement intervals were calculated from the daily frequency of bowel movements. The parameters for the models were estimated by Bayesian statistical fitting of bowel movement interval to the models. Subject MO03 was excluded from further analysis because this subject’s bowel movements consistently occurred once a day throughout the whole observation period (85 days).

To investigate individual differences, we prepared several models, some of which assumed a constant treatment effect across individuals and some assuming variation among individuals (Table S4). The models were evaluated by Widely Applicable Information Criterion (WAIC) [30]. The lower the WAIC, the better the model explains the data without overfitting. The WAIC is an index of prediction performance for unknown data. Among the models, the model considering the individual placebo effect and the test supplement effect had the lowest WAIC (Table S4). These results indicated that some subjects had increasing bowel movement frequencies as a result of intake of the test supplement; we defined these subjects as “bowel responders.”

Using the model with the best WAIC, the estimated posterior mean of each parameter and its Bayesian credible intervals were calculated for each individual (Table S5, Figure 3). The effects of the test supplement in subjects MO04, MO05 and MO10 were strong as evidenced by the 95% Bayesian credible intervals of the predicted distribution, which did not overlap zero. Therefore, MO04, MO05 and MO10 were defined as strong responders (SRs). In addition, there were 9 other subjects with estimated posterior mean values greater than zero, who were defined as weak responders (WRs). The remaining subjects were defined as nonresponders (NRs).

**Figure 3.**
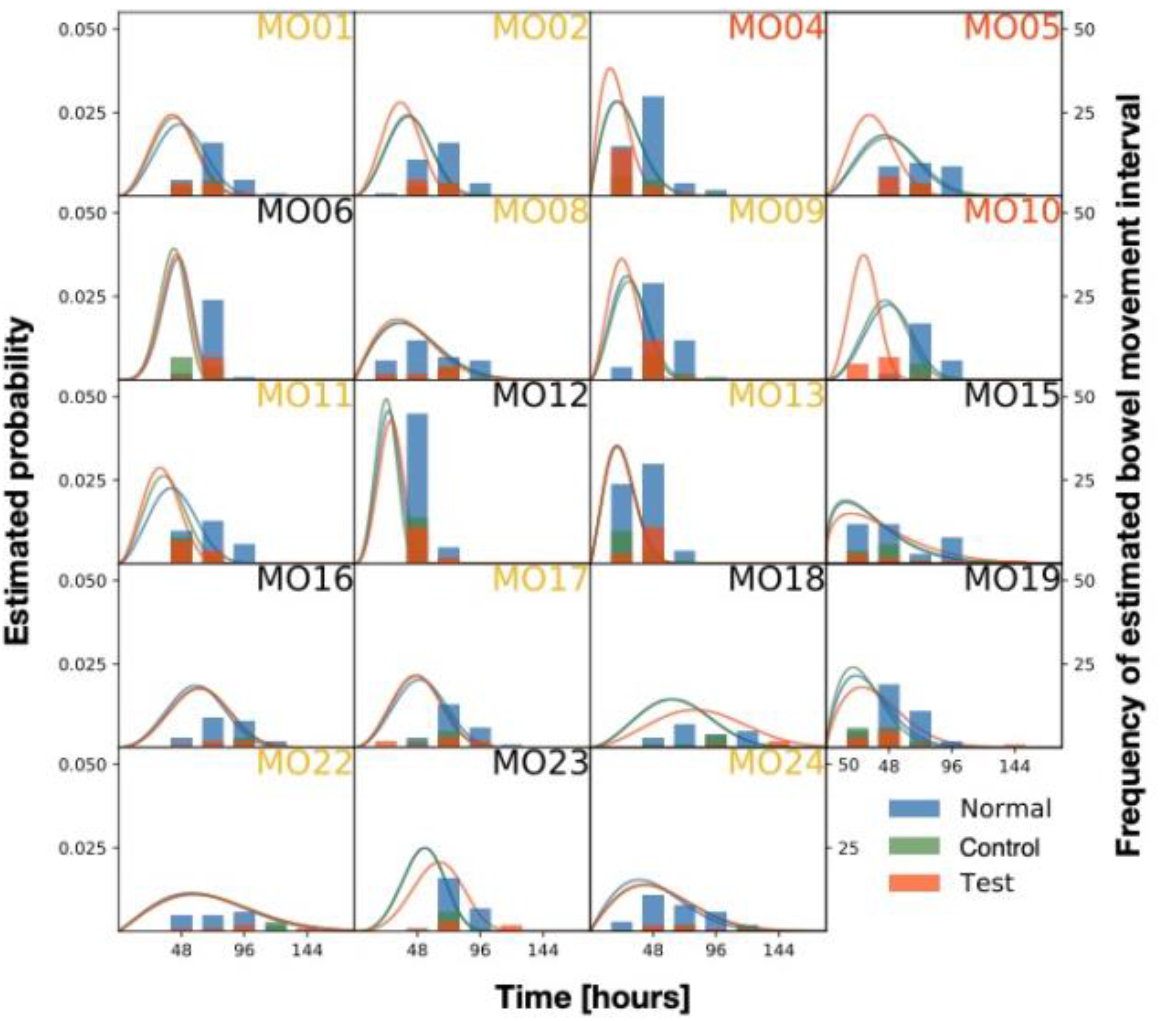
Histogram of the observed bowel movement time interval and estimated probability density function of the Weibull distribution. The vertical axis on the left shows the value of the probability density function, and the vertical axis on the right shows the frequency of the estimated bowel movement interval in the whole observation period (85 days). The responder status of the subject was defined based on the Bayesian posterior distribution: SRs are indicated in orange labels, WRs are indicated in yellow labels, and NRs are indicated in black labels.

### The abundances of fifteen genera and three metabolites differed between bowel responders and nonresponders as determined by Bayesian statistical model

We then calculated fold changes for each bacterial genus and metabolite abundance for the SR and WR groups and compared them to those for the NR group (FCs and FCw, respectively). To detect features with large or small relative abundances when the effect of the test supplement was strong, the genera and metabolites with fold change values following the order FCs > FCw > 1 or FCs < FCw < 1 were extracted, and the trends were verified using the Jonckheere-Terpstra trend test. Increasing trends were detected for five genera, and decreasing trends were detected for six genera and one metabolite (p value < 0.05) (Figure 4A). For these features, the Brunner-Munzel test was used to compare both responder groups (SRs and WRs) with the NRs group. The relative abundances of g__*Ruminococcus*_E, g__Unassigned85 (g__*Faecalibacterium* related genus), g__CAG-274 (o__Lachnospirales), g__Unassigned207 (o__Clostridiales) and g__Unassigned459 (g__*Roseburia* related genus) were significantly higher in the responder groups than in the NR group (p value < 0.05) (Figure 4B). Conversely, g__*Agathobacter*, g__CAG-24 (f__*Lachnospiraceae*), g__UBA1191 (f__*Anaerovoracaceae*), g__Unassigned66 (g__*Faecalibacterium* related genus), g__Unassigned441 (g__*Lachnoclostridium* related genus) and g__Unassigned390 (g__*Emergencia* related genus) were significantly less abundant in the responder groups than in the NRs group, and the ribulose 5-phosphate level was significantly lower in the responders (p value < 0.05) than in the NRs (Figure 4B, C). Taken together, these results revealed the bowel responders who had a bowel movement frequency-increasing effect due to *B. longum* BB536 intake and clarified that fifteen genera and three metabolites had significantly different abundances in responders compared to those in NRs.

**Figure 4.**
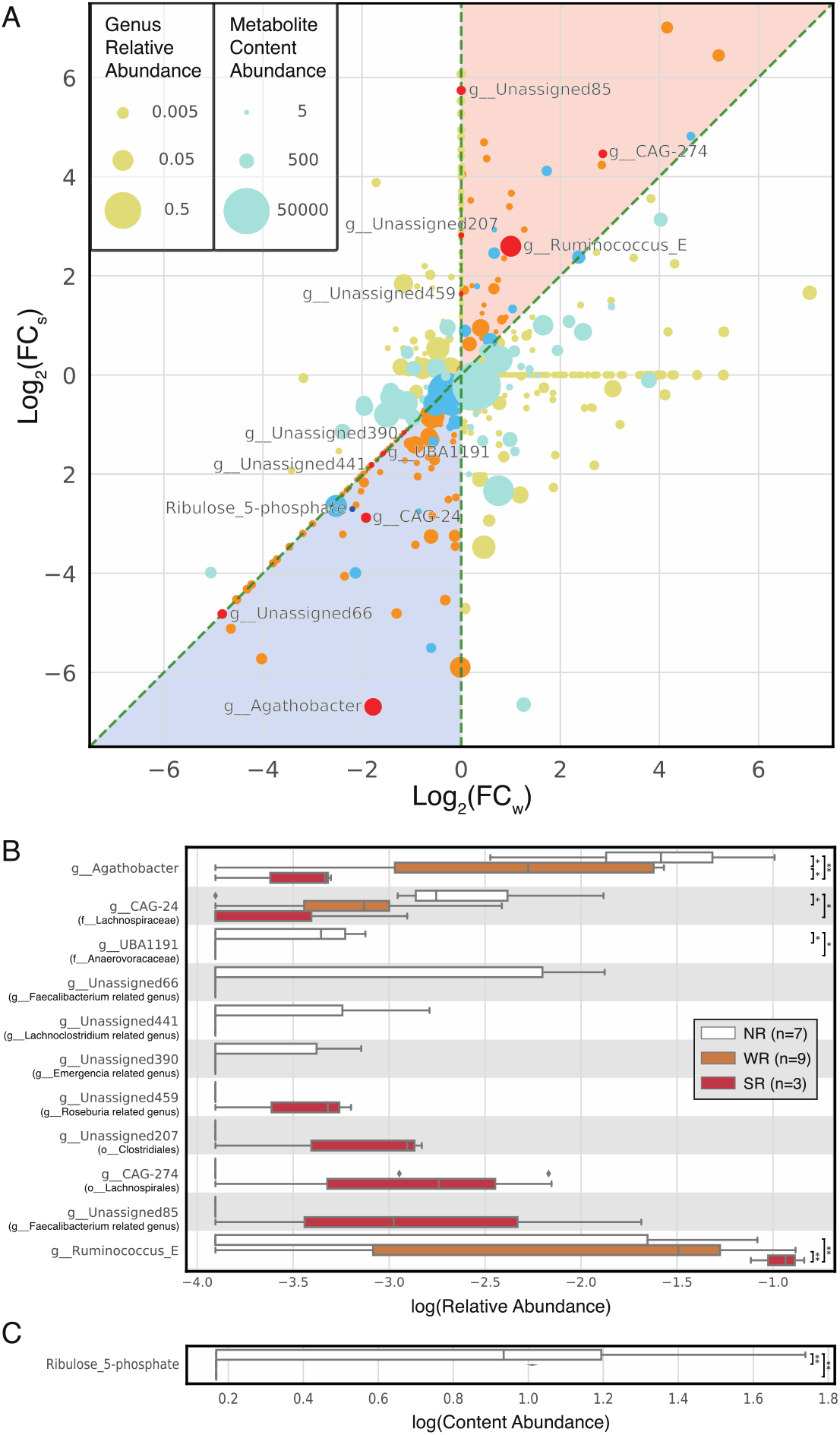
Intestinal environmental features related to the increasing bowel movement frequency due to *B. longum*B536 intake. A: A scatter plot showing the fold change in microbiota and metabolite abundances in responders compared to those in NRs. The vertical axis represents the log 2 value of the SRs’ fold change compared to that of the NRs (FCs), and the horizontal axis represents the log 2 value of the WRs’ fold change compared to that of the NRs (FCw). The warm plot corresponds to the microbiota, and its size represents the relative abundance. Red represents bacterial genera that were subjected to the Jonckheere-Terpstra trend test and had a p value < 0.05. The cold plot corresponds to metabolites, and its size represents the fecal content. Blue represents metabolites that were subjected to the Jonckheere-Terpstra trend test and had a p value < 0.05. The light orange region is FCw < FCs, that is, NR < WR < SR, and the light blue region is FCw> FCs, that is, NR > WR > SR. B, C: The relative abundance of any bacterial genus and metabolite content of fecal samples that had a p value < 0.05. Lineage names in parentheses represent the most closely related GTDB taxonomy. * indicates a p value < 0.05, according to the Brunner-Munzel test, and ** indicates a p value < 0.01. The error bars indicate the 95% confidence interval.

### Machine learning enabled us to predict bowel responders from metabologenomic data before probiotic intake

In parallel with our experimental observations of the intestinal environmental factors that determine the response to supplementation with *B. longum* BB536, we also investigated whether responders could be predicted from intestinal environmental features before *B. longum* BB536 intake by a machine learning approach. To test this strategy, we used the machine learning method combined with LASSO regression and logistic regression and predicted whether a subject would be a responder from the metabologenomic data before the test intervention (Figure 5A). The composition of bacterial genera, bacterial species, metabolites or their combinations were used as explanatory variables for responder prediction, and we conducted parameter estimation and performance evaluation in machine learning using a grid search and stratified cross validation. For the stratified cross validation, we chose 7-fold stratified cross validation to use additional training data, considering that the 7 subjects were NRs. As a result, it was possible to predict all responders (SRs and WRs), and the highest performance (area under the receiver operating characteristic curve (AUROC) = 0.810) was observed when using metabolite and bacterial genus data (Figure 5B, Table 1). Responders could also be predicted with high performance (AUROC = 0.766) using metabolite and bacterial species data, which provided more detailed information than did bacterial genera data (Table 1). LASSO regression using bacterial genus and metabolite data with a parameter set for the highest AUROC showed that the g__*Agathobacter*, g__Unassigned66 (g__*Faecalibacterium* related genus), g__*Alistipes*, g__*Mitsuokella*, g__*Parabacteroides*, g__CAG-110 (f__*Ruminococcaceae*), g__*Fusicatenibacter*, g__*Hungatella*, g__UBA1394 (f__*Ruminococcaceae*), g__CAG-65 (f__*Lachnospiraceae*), g__*Sutterella*, g__*Clostridium*_AJ, g__*Lachnospira*, g__*Sellimonas*, g__Unassigned14 (g__*Ruminococcus* related genus), g__*Agathobaculum*, g__Unassigned85 (g__*Faecalibacterium* related genus), g__*Eubacterium*_G, g__Unassigned11 (g__*Ruminococcus* related genus) and g__*Ruminococcus*_E levels were involved in the prediction of responders (Figure 5C). While the exact mechanisms underlying the B. longum-induced effects of the gut microbiota on bowel movement frequency have yet to be elucidated, our machine learning approach allowed us to predict the patients who would respond to *B. longum* BB536 intake with an increased bowel movement frequency.

**Figure 5.**
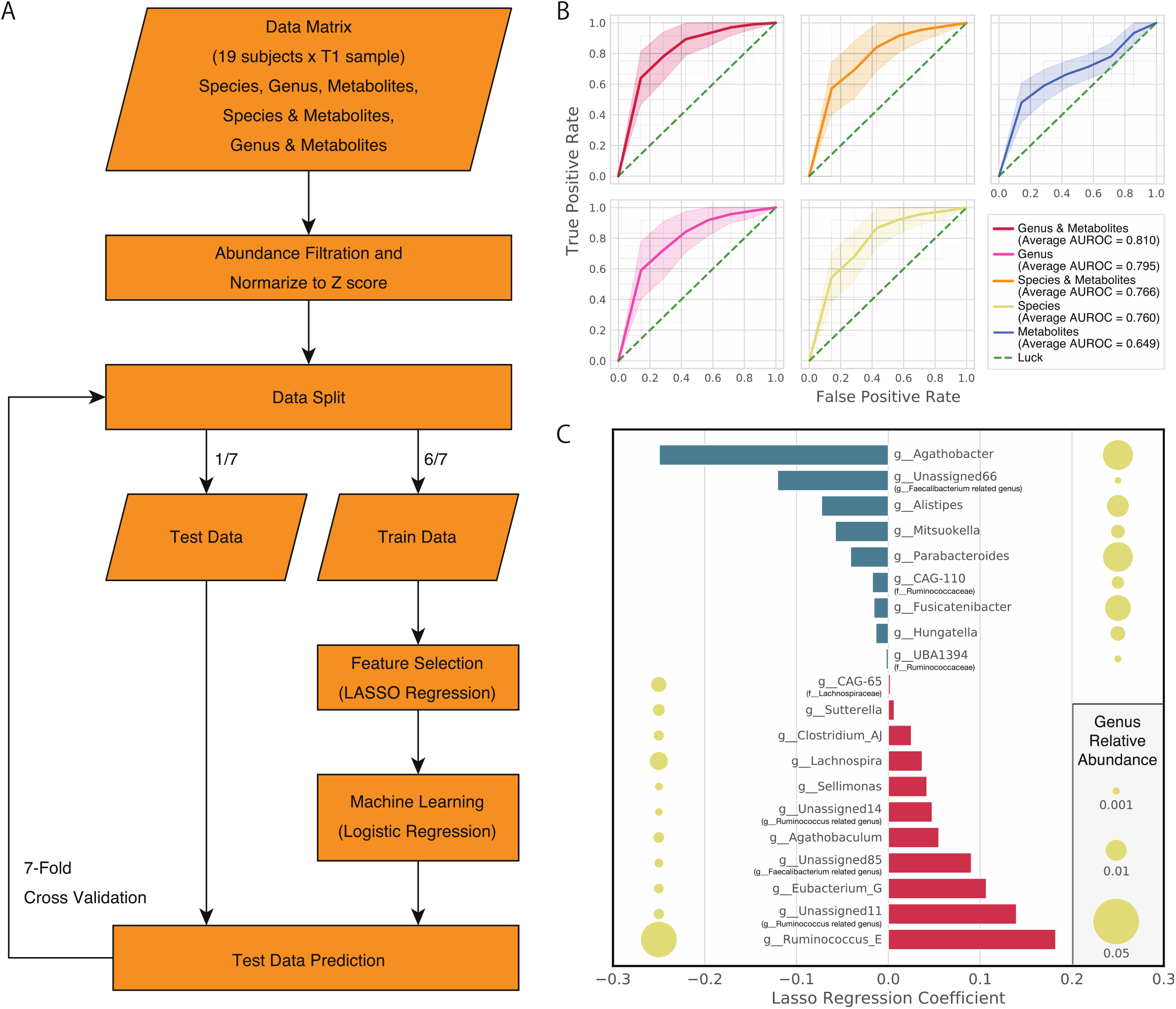
Machine learning-based responder prediction from intestinal environmental features before the intake of the test supplement. A: A flow chart of the machine learning algorithm for responder prediction. Feature selection by LASSO regression and machine learning by logistic regression algorithm were used. B: A receiver operating characteristic (ROC) curve when predicting, from intestinal omics data collected before intake of the test supplement, who will be a responder (SR and WR) (i.e., experience a significant increase in bowel movement frequency). C: Regression coefficient of the LASSO regression with respect to the responder score in the parameter with the highest accuracy in the 7-fold stratified validation, using bacterial genus and metabolite data collected before the test supplement intake began. The size of the circle adjacent to the bacterial genus name represents the average relative abundance or content (no metabolite features were detected). Lineage names in parentheses represent the most closely related GTDB taxonomy.

**Table 1.** Summary of the responder predictions from machine learning. *: AUROC; area under the receiver operating characteristic curve. **: For each data set, the parameters, AUROC, accuracy and F-measure at the highest performance are shown.

| Data | Microbiome<br>relative<br>abundance<br>threshold** | Metabolome<br>content<br>abundance<br>threshold** | LASSO<br>regression<br>parameter<br>alpha** | Average AUROC | Accuracy | F-measure |
| --- | --- | --- | --- | --- | --- | --- |
| Genus and<br>metabolite | 0.001 | 10000 | 0.001 | 0.810 | 0.746 | 0.797 |
| Genus | 0.001 | - | 0.005 | 0.795 | 0.739 | 0.797 |
| Species and<br>metabolite | 0.005 | 10000 | 0.1 | 0.766 | 0.724 | 0.788 |
| Species | 0.005 | - | 0.1 | 0.760 | 0.721 | 0.787 |
| Metabolite | - | 100 | 0.001 | 0.649 | 0.617 | 0.675 |

## Discussion

The bowel movement responses to probiotic intake vary among individuals [11], and the gut microbiota is a key factor that can account for these differences [17]. Thus, in this study, we accurately quantified individual differences in the increase in bowel movement frequency in response to intake of *B. longum* BB536 contained in acid-resistant seamless capsules by a statistical model and defined the responders as those whose bowel movement frequency increased noticeably. In addition, the machine learning analysis revealed that responders could be predicted from features of the intestinal environment before intake of the test supplement began, and the predictive performance was improved by using both microbiome and metabolome data. These results suggest that accurate quantification of individual response intensity and machine learning predictions may enable companion diagnostics for response to diet based on the intestinal environment.

Our finding that using both microbiome and metabolome data improved the predictive performance of machine learning shows that a metabologenomic approach combining metagenomics and metabolomics may be important for analyzing the response depending on the individual intestinal environment. In recent years, multiple studies have predicted responses to intervention from metagenomic datasets alone, including datasets of taxonomic composition [22,23,31]; incorporating metabolome data may enhance predictive performance. In addition, previous studies have reported that, in addition to increasing bowel movement frequency, the intake of the *B. longum* BB536 strain encapsulated in an acid-resistant seamless capsule results in a fecal color change from dark to light, a fecal shape change from a hard or watery state to a soft state and a decrease in the amount of ammonia in feces [14,32]. If individual responses with these effects were accurately quantified, then metabologenomic data may be able to predict which subjects will have these responses, as well as which will have increased bowel movement frequency. In addition, several studies have suggested that individual responses to the intake of other dietary factors could be predicted from the intestinal environment [20,21,33]. From these cases, it is expected that it will be possible to predict the effect of probiotics from the intestinal environment before intake in the future, and the development of personalized health care and medical businesses targeting the intestinal environment are expected by using this prediction system.

We established an accurate Bayesian statistical model to estimate the bowel movement frequency-increasing effect due to *B. longum* BB536 intake, and this model revealed which subjects were responders or NRs in this study. This model enabled us to detect shortening of the bowel movement interval due to the intake of *B. longum* BB536, which is consistent with previous research [15,16]. In addition, because Bayesian estimation of the parameters enabled us to calculate a correct interval, this model achieved accurate quantitative evaluation of the effect of *B. longum* BB536 intake on bowel movement frequency, which has been difficult to achieve by threshold-based responder estimation [21].

Few significant differences in the microbiome and metabolome were detected among the baseline, *B. longum* BB536 intake and control supplement samples. This lack of significant differences in the intestinal environment is related to interindividual variation, which masks the effect of the intake of *B. longum* BB536. This finding has also been reported in several studies that have observed changes in the gut microbiome composition by probiotics or diets. Thus, this effect is because the intestinal environment of each individual was robust and individual differences in the intestinal environment override the effect of probiotic intake.

Our observational and machine learning results indicated that a higher or lower relative abundance of 27 genera was associated with whether a subject responded well to *B. longum* BB536 intake, and intriguingly, 23 of these 27 responder-related genera belonged to the class Clostridia. Clostridia is an important bacterial group that produces mainly short-chain fatty acids (SCFAs), including butyrate, in the gut environment [34]. An imbalance of SCFAs may cause IBD and IBS because of the involvement of SCFAs in the maintenance of intestinal homeostasis [35–37]. Clostridial clusters IV and XIVa in Clostridia are reported to produce butyrate, which is related to the promotion of large intestinal peristaltic movement [34]. Among the 23 genera suggested as possibly involved in the bowel responders in this study, nine *Ruminococcaceae* bacteria belong to Clostridial cluster IV, and ten *Lachnospiraceae* bacteria belong to Clostridial cluster XIVa. The bacteria in Clostridial clusters IV and XIVa cross-feed with *Bifidobacterium* and metabolize the inulin-type fructans derived from *Bifidobacterium* to produce butyrate [38–40]. We therefore suggest that the balance of bacteria belonging to Clostridial clusters IV and XIVa may be related to the increase in bowel movement frequency in response to intake of *B. longum* BB536 via butyrate or other SCFAs and that researching the cross-feeding relationship of these bacteria might lead to a detailed understanding of this relationship.

Our results also suggest that butyrate may play a role in the response to *B. longum* BB536 intake. Compared with NRs, responders tended to show an increase in the abundance of several butyrate-producing bacteria during the test intervention period (Figure 4). This result also supports a relationship among the gut microbiota, butyrate and an increase in bowel movement. Furthermore, it has been reported that the efficiency of butyrate production in the sugar metabolism of these bacteria varies among bacterial species and habitat. Previous studies have indicated that the sugar-metabolizing abilities of *Ruminococcus bromii*, *Eubacterium rectale* (*Agathobactor rectale*), *Bacteroides thetaiotaomicron*, *Bifidobacterium adolescentis*, *Anaerostipes caccae* and *Eubacterium hallii* vary with habitat [39,41]. In addition, *Ruminococcus* and *Bifidobacterium* are localized in the intestinal lumen, and *Clostridium* is localized in the intestinal lumen and on the intestinal epithelial cell surface, whereas *E. rectale* preferentially colonizes the mucous layer to produce and promote the utilization of butyrate by colonic epithelial cells [34,42]. Taken together, these findings suggest that variation in butyrate productivity among bacteria and their locations in the large intestine may influence whether a subject responds to *B. longum* BB536 intake.

In our study, the most valuable points are the clinical trial with a double-blind placebo-controlled design and multi-omics features to define responsiveness to probiotic intervention; however, two limitation points should be considered. First, 16S metagenomic data were used to evaluate the gut microbiome. Since 16S metagenomic data reveal only the taxonomic composition and cannot be used for detailed functional analysis, it is necessary to conduct a future study with shotgun metagenomics to clarify the hypothesis proposed in this study. The other limitation is that the final number of subjects was 20. In this study, acquiring the time-series data of each individual compensates for that; however, in the future, the results need to be validated by a larger cohort study. Although no such large cohort has been conducted for the effect of probiotics on bowel movement frequency, the expectations and demands for such a study have risen considerably.

## Conclusions

In summary, our results show that the gut microbiota composition has a strong impact on the effectiveness of *B. longum* BB536 supplementation in increasing bowel movement frequency, and the gut metabologenomic data enable us to predict the effect of *B. longum* BB536 supplementation before intake. These findings have interesting implications for personalized treatment of chronic constipation. In addition, these findings suggest that the influence of the intestinal environment on the response to intervention should be considered not only for probiotics but also for all health effects, adverse effects and side effects for all oral intake interventions. When the effect of probiotics, diets or drugs is dependent on the intestinal environment before intake, as it is in the present study, the demand for intestinal environment alterations to enhance the effect may also increase. Furthermore, the development of personalized health care and medical businesses targeting the intestinal environment are expected to use this intervention effect quantification and responder prediction system.

## Methods

### Trial design and recruitment

In this study, we conducted a randomized double-blind controlled crossover trial with healthy subjects from September to December 2015 to quantify the increase in bowel movement frequency in response to intake of acid-resistant seamless capsules containing *Bifidobacterium longum* BB536. In addition, we analyzed the microbiome and metabolome profiles of fecal samples collected during the trial period (Figure 1A, Figure S4, Table S6) (see also Supplementary Protocol).

The sample size was estimated based on a previous randomized controlled trial using the probiotic strain *B. longum* BB536 as the test supplement [43]. In this previous trial, the bowel movement frequency during the test intervention period was 9.6 ± 0.6 [/ 2 weeks], and that during the control intervention period was 8.4 ± 0.4 [/ 2 weeks]. We applied power analysis to estimate the number of subjects with a significance level of 5%. The statistical power was 80%, with the expectation that 20% of subjects would drop out in this trial. From these conditions, the number of participants required for this trial was estimated to be eight.

A preliminary bowel movement test was conducted for 50 Japanese participants, and 24 subjects who fulfilled the criteria for age, male-female ratio and bowel movement status were selected before the main trial from the preliminary results. Four out of 24 subjects were not used in the subsequent analysis, as they violated the protocols (by taking medication or failing to collect fecal samples). In these twenty subjects, the average age was 47.7 ± 5.8, and the male/female ratio was 8:12. The remaining baseline demographic and clinical characteristics did not significantly differ between the control and treatment groups (Table S7).

### Subject characteristics and exclusion criteria

This clinical study was approved by the ethics committee and then requested to CPCC Co., Ltd. All of the participants consented to participate in this study after the study was fully explained to them.

The subject inclusion criteria were as follows: 1) aged 40 to 60 years at the time of providing consent and 2) stool frequency of three to five times per week or more than seven times per week. The exclusion criteria were as follows: 1) laparotomy surgery performed within six months before the start of the study, 2) intake of antibiotics for more than one week within six months before the start of the study, 3) allergies to the test supplement, 4) significant lifestyle changes intended during the examination period, 5) susceptibility to chronic diarrhea, 6) history of significant liver dysfunction, gastric dysfunction, or cardiovascular disease, 7) suspected chronic or acute infection, 8) known or potential pregnancy or breastfeeding, 9) participation in other trials within the month before the start of the study and 10) otherwise assessed to be inappropriate for the study by the physician involved in the study. All subjects were monitored for defecation frequency for two weeks before the start of the study, and it was confirmed that the defecation frequency was either three to five times per week or seven times per week or more. Among the subjects who completed the study, those who met any of the following conditions were excluded from further analysis: 1) considered to have met one or more of the exclusion criteria, 2) an inoculation rate of the test supplement less than 80%, 3) large variation in the diet record or lack of confirmation of no change from the record, 4) large variation in the content of the lifestyle record or lack of confirmation of no change from the record, 5) continued or repeated ingestion of medicines, foods or supplements that might influence the outcomes of the trial and 6) otherwise assessed to be inappropriate for the study.

### Trial intervention: dietary information

In this trial, we used acid-resistant seamless capsules containing approximately 5.0 * 10_9_ *B. longum* BB536 (0.53 g per serving) as the test supplement (MORISHITA JINTAN CO., LTD, Japan). These capsules are spherical with a diameter of approximately 2.4 mm and consist of two types of acid-resistant layers. It has been found that approximately 90% of encapsulated *B. longum* BB536 survive for two hours in artificial gastric fluid adjusted to pH 1.2 [12]. The acid-resistant pH-dependent disintegrating membrane, which is the outermost layer of the capsule, passes through the stomach and collapses as the surrounding pH becomes neutral in the small intestine. Subsequently, the hardened fat layer, which is an intermediate layer, is dissolved by the surface-active action of bile acid, the digestive action of lipase and physical stimulation by intestinal motility, and then the internal bacteria are released. The released bacteria have been reported to grow in the intestine [44,45]. For the control supplement, the same capsule containing only starch was used (MORISHITA JINTAN CO., LTD, Japan). The energy, protein, lipid, carbohydrate, and sodium contents were adjusted to be equivalent between the test supplement and the control supplement. In addition, the appearances of the test and control supplements were processed to be indistinguishable.

### Trial intervention: randomization and blinding

Randomization in the trial was performed by a blocked stratified randomization method. First, the 24 subjects who passed the selection criteria were assigned to two groups (group A, group B) by stratification, taking into consideration age, male-female ratio and bowel movement status before the trial period. Subsequently, the symbol “A” or “1”, representing either the test or control supplement, respectively, was randomly assigned to each group of subjects, and a test supplement assignment table with the test supplement symbol and the subject identification code was prepared. Immediately after the test supplement was assigned to one group, the table was sealed and maintained secure until the end of the study such that most of the investigators were blinded to the group assignments. Each subject orally took one serving of the test or control supplement per day with as much water as they desired. One kind of intervention was taken for two weeks, and then the other kind was taken for two weeks after a four-week washout period. The capsules were stored at room temperature by the subjects until intake.

### Trial outcomes and sample collection

The primary outcomes in this study were bowel movement frequency and the features of the intestinal environment (intestinal microbiome and metabolome). The bowel movement frequency data as well as data on other activities were acquired by a questionnaire that was completed once a day (Table S8). The questionnaire included life status, diet and bowel movement status. All subjects were prohibited from consuming 1) beverages or foods containing lactic acid bacteria or bifidobacteria in large amounts, 2) beverages or foods rich in dietary fiber or oligosaccharides, 3) dietary supplements, 4) functional yoghurt, and 5) fermented soybeans (natto), and each participant noted on the questionnaire if any of these items had been consumed. The features of the intestinal environment were obtained from the stool specimen collected one week before the intervention, during the second week of the intervention period, and one week after the intervention. Feces were collected by each subject at home using a collection sheet (Nagaseru; OZAX, Japan) laid in the toilet bowl and a feces collection tube (feces container 54 × 28 mm; SARSTEDT AG & Co, Germany). After collection, the stool samples were stored immediately in the household freezer and then collected later and transported to the laboratory by refrigerated transport.

### DNA extraction and 16S rRNA gene analysis

We used the protocol described in a previous report [46] for DNA extraction from stool samples. From the extracted DNA samples, the 16S rRNA gene region was amplified using the universal primers 27Fmod and 338R for the V1-V2 region of the bacterial 16S rRNA gene [47]. For sequencing of the amplicon DNA, a MiSeq platform (Illumina, USA) was used in paired-end mode with 600 cycles. The obtained 16S rRNA gene sequence data are available in the DDBJ DRA (DRA accession number: DRA006874).

For 16S rRNA gene analysis, the sequenced forward and reverse reads of each sample (average number of reads: 44,441 ± 5,959) were merged using vsearch version 1.9.3 (options: --fastq_maxee 9.0 --fastq_truncqual 7 --fastq_maxdiffs 300 --fastq_maxmergelen 330 --fastq_minmergelen 280) [48]. Subsequently, we obtained high-quality reads by quality filtering using fastp with the default parameters [49]. High-quality read FASTQ files were converted to FASTA format using SeqKit [50]. The final set of high-quality reads was used for identifying exact sequence variants (ESVs) using the Deblur pipeline [51]. In the pipeline, artefact sequences including PhiX were removed, and de novo chimaera sequences were analyzed and removed. All ESVs were aligned to the Genome Taxonomy Database (GTDB) version 86 [52] using BLAST version 2.8.1 [53], and 99% identity was used as the threshold for the species level [54], whereas 94% identity was used for the genus level [55]. The correspondence table of ESVs and phylogenetic information and the number of reads in each step are shown in Table S9 and S10.

### Metabolite extraction and analysis

Metabolome analysis was conducted as described previously with some modifications (PMID: 25525179, 26798400). To extract metabolites from feces, samples were lyophilized using a VD-800R lyophilizer (TAITEC) for at least 24 h. Freeze-dried feces were disrupted with 3.0 mm zirconia beads by vigorous shaking (1,500 rpm for 10 min) using a Shake Master NEO homogenizer (Biomedical Science). A total of 500 μl of methanol containing the internal standards (20 μ_M_ each of methionine sulfone and D-camphor-10-sulfonic acid (CSA)) was added to 10 mg of disrupted feces. Samples were further disrupted with 0.1 mm zirconia/silica beads by vigorous shaking (1,500 rpm for 5 min) using the Shake Master neo. Next, 200 μl of ultrapure water and 500 μl of chloroform were added before centrifuging at 4,600×g for 15 min at 20°C. Subsequently, 150 μl of the aqueous layer was transferred to a centrifugal filter tube (UltrafreeMC-PLHCC 250/pk for Metabolome Analysis, Human Metabolome Technologies) to remove protein and lipid molecules. The filtrate was centrifugally concentrated and dissolved in 50 μl of ultrapure water immediately before CE-TOFMS analysis. The measurement of extracted metabolites in both positive and negative modes was performed by CE-TOFMS. All CE-TOFMS experiments were performed using the Agilent CE capillary electrophoresis system (Agilent Technologies). Annotation tables were produced from measurements of standard compounds and were aligned with the datasets according to similar values and normalized migration times. Then, peak areas were normalized against those of the internal standards, which were methionine sulfone and CSA for cationic and anionic metabolites, respectively. Concentrations of each metabolite were calculated based on the relative peak areas and the concentrations of standard compounds. The obtained metabolome data are available in Table S11 and 12.

### Bioinformatics and statistical analyses

All statistical analyses were performed using Python. The Shannon diversity index was used for alpha diversity, and the Spearman correlation distance was used for beta diversity. MDS was calculated using beta diversity. A two-sided Wilcoxon signed rank test was used to compare bowel movement frequency data between the two groups. A two-sided Brunner-Munzel test was used for comparing metabologenomic data between the two groups. A two-sided Jonckheere-Terpstra trend test was used to verify the trend in metabologenomic data among the groups. The Benjamini-Hochberg false discovery rate correction (FDR) method was used for multiple test correction. For gut microbiome analysis, taxonomic composition data in the genus, species, and ESV levels were used. Of these, ESV-level data were used only to calculate the alpha diversity. The genus-level data were used for the other statistical analyses. The species-level data were used only for the prediction of bowel responders. For gut metabolome analysis, relative area compared with that of an internal standard and content of metabolite data was used. The metabolite relative area data were used only to calculate the alpha diversity. The metabolite content data were used for the other statistical analyses.

### Bowel responder definition based on Bayesian Weibull regression

Using the data from the questionnaires recorded by each subject over the course the study, subjects who strongly responded to treatment, i.e., who showed an increased bowel movement frequency, were estimated using a statistical model. An increasing effect of the test supplement on bowel movement frequency was defined as a shortened interval between bowel movements; however, other measures of bowel movement frequency or bowel movement probability were also evaluated.

Paying attention to the fact that bowel movement tends to occur with the lapse of time, the bowel movement interval of each individual was modeled by the Weibull distribution. The Weibull distribution is widely used to investigate the effects of drugs on survival time, and it has a shape parameter and a scale parameter [56]. The effect of intake itself was estimated based on a proportional hazards model, assuming that intake influences the scale parameter. The corresponding mathematical formulas are shown below.

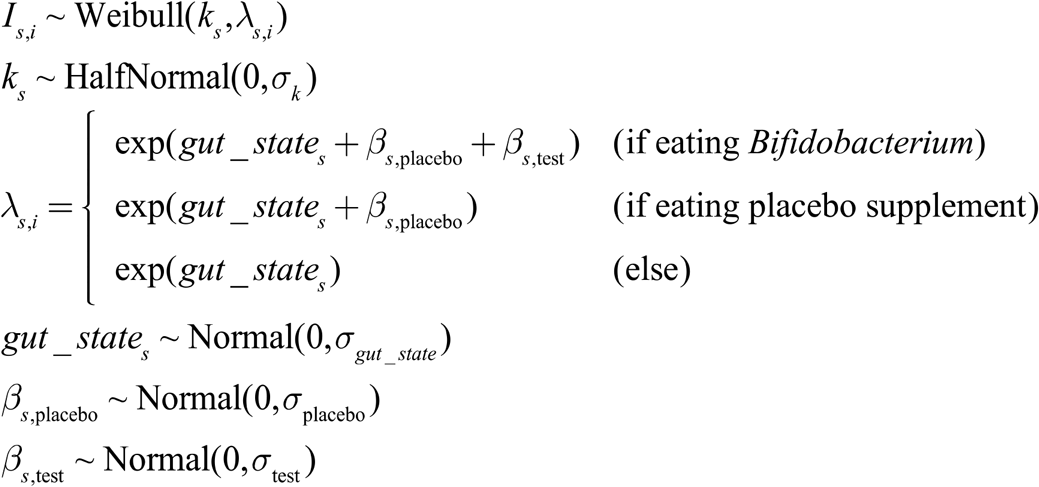

*s*: subject index
*i*: stool index

*I_s_,_i_*: time interval of *i*th stool of subject *s*

*k_s_*: shape parameter of subject *s*

*λ_s_*,*_i_*: scale parameter of *i*th stool of subject *s*

*σ_k_*: scale parameter of shape parameter *k*

*gut*_*state_s_*: personal variable parameter to explain normal gut status

*β_s_*_,placebo_: personal variable parameter to explain intake effect occur in both *Bifidobacterium* and controll supplement

*β_s_*_,test_: personal variable parameter to explain intake effect occur in only *Bifidobacterium* supplement

*gut*_*state_s_*: personal variable parameter to explain normal gut status

*σ_gut_state_*: scale paramter of *gut_state*

*σ*_placebo_: scale paramter of *β_s_*_,placebo_

*σ*_test_: scale paramter of *β_s_*_,test_

In this model, it is assumed that the effect of each variable at time t occurs as a product of the hazard function in the basis state. The hazard function is a function that expresses the probability of occurrence of an event after time lapse Δt from time t. In this study, it was assumed that the effect of the variable accompanying the bowel movement time, such as the influence of the intake of the test supplement, occurs as a product of the hazard function in the normal state and that the occurrence probability of bowel movement changes. This assumption is equivalent to the assumption that factors other than intake do not influence the hazard function throughout the observation period. This assumption is supported by the empirical observation that the condition in the intestine is stable in a state where food intake is strictly restricted, as it is in the cohort of the present study. Moreover, the parameters in the Weibull distribution in the model and the effect of intake are unknown. Such values were estimated from the observed data. The record of bowel movements acquired in this study did not include the time of each movement but reported the number of bowel movements per day. Therefore, the interval between successive bowel movements was estimated by dividing 24 h by the frequency. One subject (MO03) consistently had one bowel movement a day throughout the entire observation period of 85 days; thus, the bowel movement interval was always 24 h. Since a variable with zero variance is inappropriate for model estimation, this subject was excluded from subsequent analysis. Furthermore, multiple models were constructed and compared using the Widely Applicable Information Criterion (WAIC) [30]. WAIC approximates the prediction error (generalization error) for unknown data of the model; WAIC can also be used for a model in which the posterior distribution of parameters cannot be approximated by a normal distribution [57].

Parameter estimation was performed by a Hamiltonian Monte Carlo method based on NUTS algorithm using Python and Stan [58]. A generalized linear model of the Weibull distribution using Markov Chain Monte Carlo (MCMC) was applied following a previous study [59]. The number MCMC iterations was 20,000, the number of chains was 8, and the first 1,000 steps in each iteration were discarded as burn-in. In a previous study, MCMC convergence was confirmed by a potential scale reduction factor (PSRF or Rhat) less than 1.1 for all estimates [60]. Similarly, in the present study, Rhat was calculated based on the variance of multiple MCMC chains and was used for convergence determination. Initial values were used for the other parameters. The implemented software is available from https://github.com/metagen/EEBIIC.

### Other defecation responder estimation models

The defecation dynamics were modeled as defecation frequency and defecation probability. Regarding defecation frequency, it was assumed that the number of defecations per day for each individual followed a Poisson distribution. The Poisson distribution is used when modeling count data and has one ratio parameter. The effect of ingestion was estimated assuming that the ratio parameter changes according to ingestion. For this method, the questionnaire-based defecation frequencies obtained from each subject were used.

Regarding defecation probability, it was assumed that occurrence of defecation per day of each individual can be modeled by the Bernoulli distribution. The Bernoulli distribution is used in expressing the occurrence probability of alternative choices, such as the side of a coin, and has one probability parameter. The effect of ingestion was estimated assuming that the probability parameter changes through a logistic function according to ingestion. For this method, the defecation frequency data were converted to presence/absence data of defecation each day, with 1 recorded if a bowel movement occurred and 0 otherwise.

### Prediction of responders by machine learning

We used the relative abundance of bacterial species or genera and the fecal metabolite content before the test intervention (T1) of 19 subjects, i.e., all subjects except MO03, as feature values. MO03 was excluded from this analysis because the bowel movements of this subject always occurred once a day throughout the whole observation period (85 days), which was not suitable for determining responders by the Bayesian statistical model. Of all feature values, those of the relative abundance or the fecal content exceeding a certain threshold were selected. For the threshold value, a grid search was conducted to search for the one with the maximum predicted AUROC. Subsequently, each feature value was standardized by centering to a mean of 0 and dividing by the standard deviation (z-score) of each feature, considering the following potential problems: 1) it is possible that a feature value with a large range of possible values will have a large influence on the prediction because the range of values that can be taken with the microbial relative abundance and metabolite contents are different; and 2) the feature importance of the microbiome and metabolome cannot be directly compared. Nineteen subjects were classified into two groups, responders (SRs and WRs) and NRs. To identify metabologenomic markers that can distinguish responders from NRs, we constructed classification models based on metabologenomic data using LASSO regression and logistic regression algorithms. The model was validated by 7-fold stratified cross-validation testing (we resampled dataset partitions 100 times). A LASSO regression model was used for feature selection from the training dataset, which was fitted to the mean value of the increase in bowel movement frequency with the test supplement for each individual estimated by a statistical model. Only the features with nonzero coefficients were extracted. Regarding the LASSO regression hyperparameters, a grid search was performed to search for the parameter set with the maximum predicted AUROC. Test data were predicted by a logistic regression model that learned from feature-selected training data. For a hyperparameter in logistic regression, we used the default parameters in sklearn.linear_model.LogisticRegression (version 0.19.1).

Z-score-standardized features, which were bacterial genera of relative abundance > 0.001 or metabolites present at > 10,000 [nmol/g] in fecal content before test supplement intake and were the conditions with the highest prediction AUROC values in the cross validation, were used for LASSO regression (alpha = 0.001). As a result, the regression coefficients of each bacterial genus and metabolite were calculated, and the features contributing to the responder prediction were extracted.

## Data Availability

The raw sequencing dataset reported in this paper is available in the DDBJ Sequence Read Archive (DRA) repository (DRA006874; https://ddbj.nig.ac.jp/DRASearch/submission?acc=DRA006874). The metabolomic datasets by CE-TOFMS are included within the article and its additional files.

## List of abbreviations

IBS: irritable bowel syndrome
IBD: inflammatory bowel disease
MDS: multidimensional scaling
FDR: false discovery rate
WAIC: Widely Applicable Information Criterion
SRs: strong responders
WRs: weak responders
NRs: nonresponders
AUROC: area under the receiver operating characteristic curve
SCFAs: short-chain fatty acids
ESVs: exact sequence variants
CSA: D-camphor-10-sulfonic acid
MCMC: Markov Chain Monte Carlo
PSRF: potential scale reduction factor

## Declations

### Ethics approval of research

The human rights of the subjects who participated in this study were protected at all times, and the study observed the Helsinki Declaration and Ethical Guidelines on Epidemiological Research in Japan referring to cases concerning standards for clinical trials of drugs. This randomized controlled trial was conducted with the approval of the clinical trial ethics review committee of Chiyoda Paramedical Care Clinic.

### Patient consent

Obtained.

### Clinical trial registration

UMIN-CTR, UMIN000018924. Registered 07 September 2015, https://upload.umin.ac.jp/cgi-open-bin/ctr_e/ctr_view.cgi?recptno=R000021894

### Competing interests

The authors declare no conflict of interests.

### Funding

None declared.

### Author contributions

S.F. and T.Y. conceived and designed the project. M.K., T.Y. and S.F. designed the clinical trial. M.K. prepared the test supplement used for intervention. S.M., C.I. and Y.I. extracted DNA from fecal samples. C.I. and Y.I. performed the microbiome analysis. S.M., C.I. and Y.I. performed the CE-TOFMS-based metabolome analysis. Yuya N. conducted statistical analysis. All analyses related to metabologenomic data were performed by Yuya N. Shinya S. performed responder classification by Bayesian modeling. Yuya N. predicted responders from metabologenomic data by machine learning. Y.M. performed the PCR amplification of the 16S rRNA region of *Bifidobacterium*. Yuichiro N., N.W., K.H. and J.U. contributed to the study design. Yuya N. and Shinya S. constructed figures and described the thesis. All authors reviewed and approved the final version of the paper.

## Acknowledgments

We acknowledge Shuji Suzuki for his insight and feedback regarding responder prediction by machine learning. We would like to thank MORISHITA JINTAN CO., LTD, and CPCC Co., Ltd., who conducted the clinical trial. We also thank the Tokyo Institute of Technology Yamada laboratory and Metabologenomics, Inc., staff, who participated in discussion of this research.

## Authors’ information

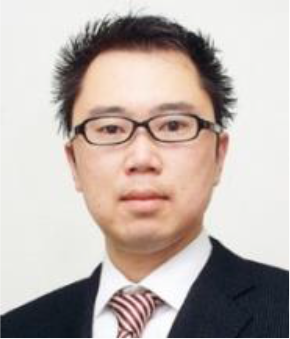

Name

Dr. Takuji Yamada

## Biography

Dr.Takuji Yamada received his doctorate (Doctor of Science) in 2007 from Kyoto University. That year, he was appointed Assistant Professor at the Institute for Chemical Research, Kyoto University. He was then a postdoctoral fellow (2008-2010) and a technical officer (2010-2012) at the European Molecular Biology Laboratory (EMBL). In 2012, he joined Tokyo Institute of Technology as Associate Professor in the Department of Biological Information. Since 2016, he has been Associate Professor in the Department of Life Science and Technology, School of Life Science and Technology, Tokyo Institute of Technology.

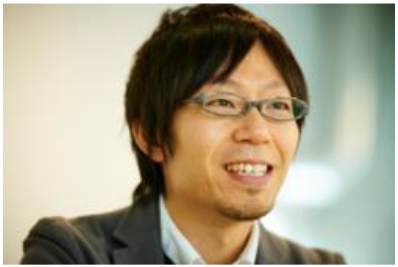

Name

Dr. Shinji Fukuda

## Biography

After obtaining his Ph. D. (Gut Microbiology) at Graduate School of Agriculture, Meiji University in 2006, Dr. Shinji Fukuda undertook several research positions at RIKEN. He was then appointed to Project Professor of the Institute for Advanced Biosciences, Keio University in 2019. Currently, he is also holding other posts as visiting professor in University of Tsukuba School of Medicine from 2016; group leader of Intestinal Ecosystem Regulation Group in KISTEC-KAST from 2017; Co-PI of ERATO in JST from 2019; and visiting professor in University of Technology Malaysia from 2019. He founded and is President CEO of Metabologenomics, Inc. from 2015.

---

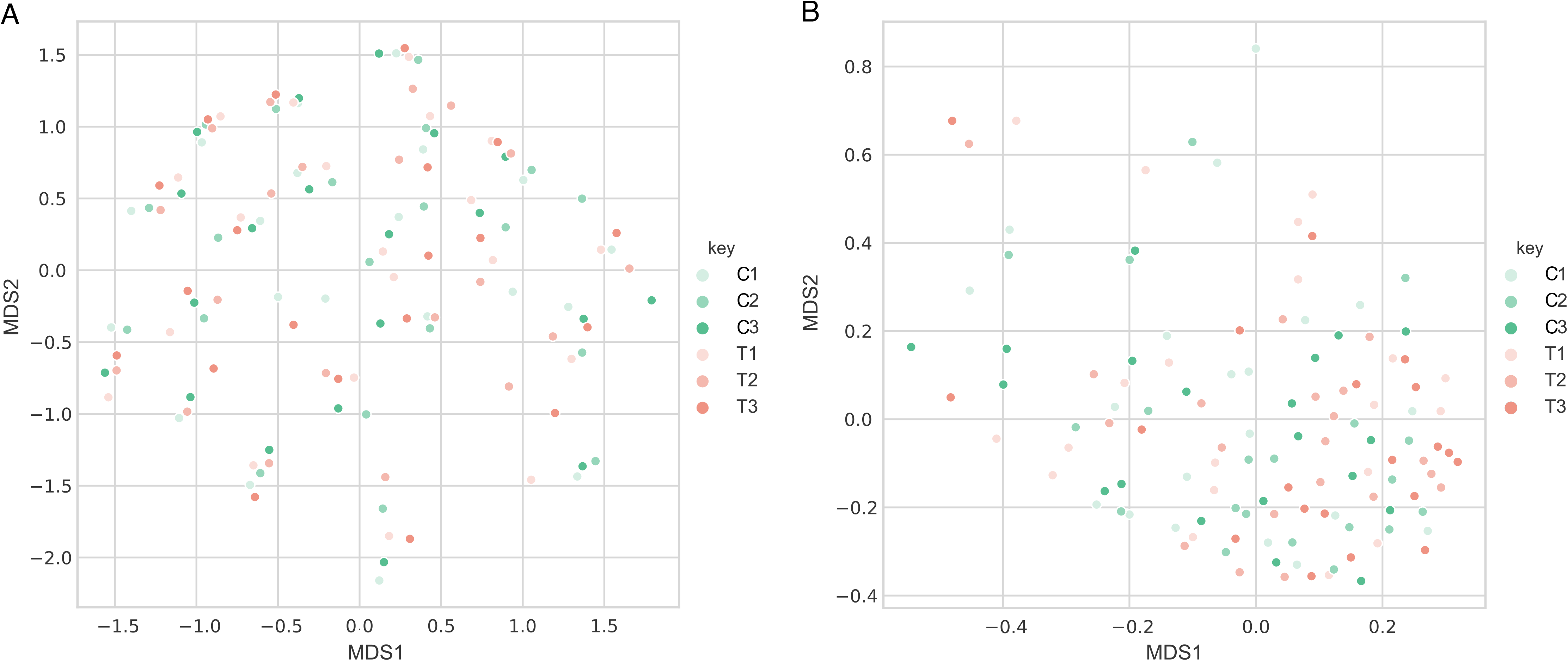

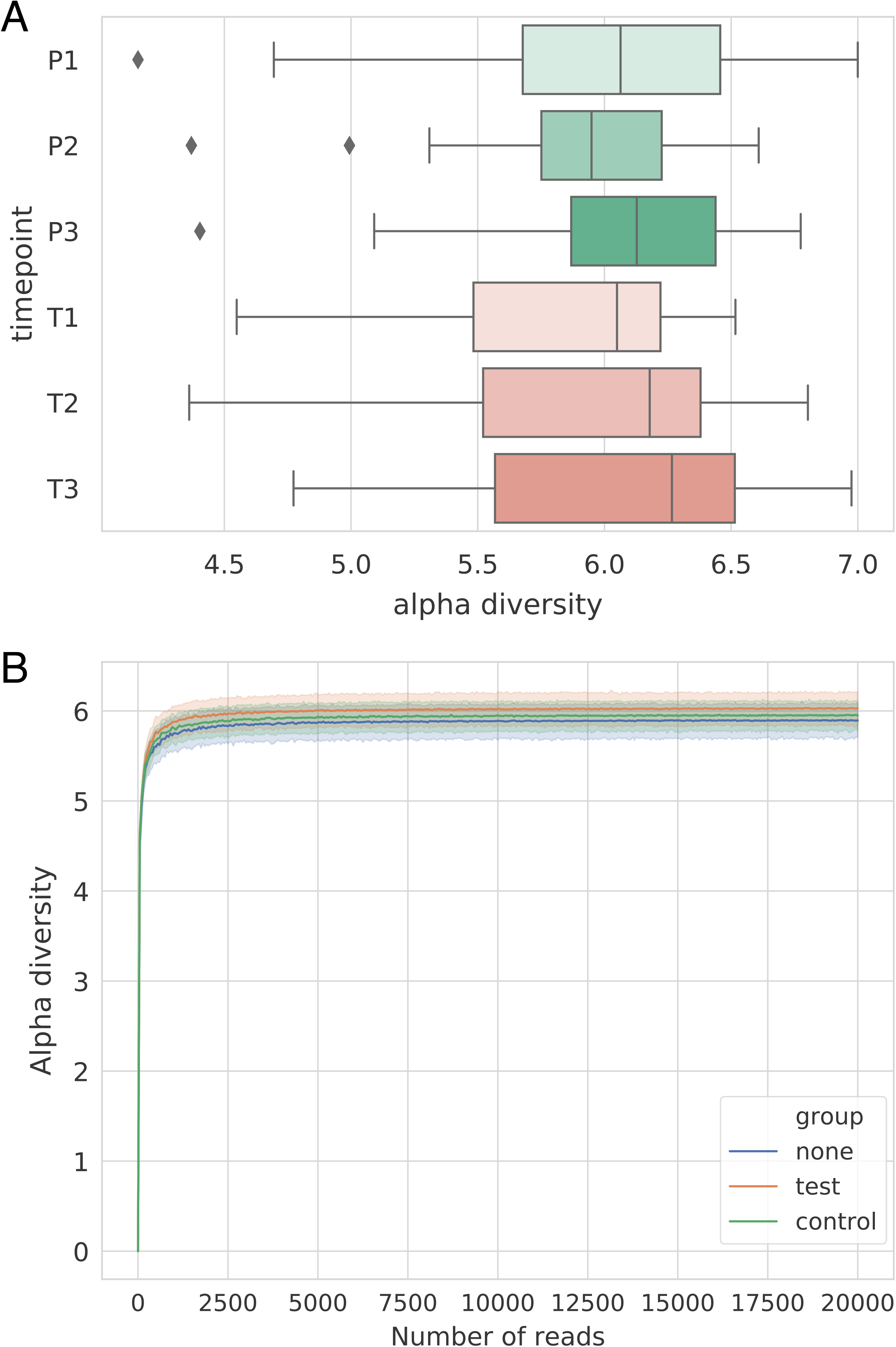

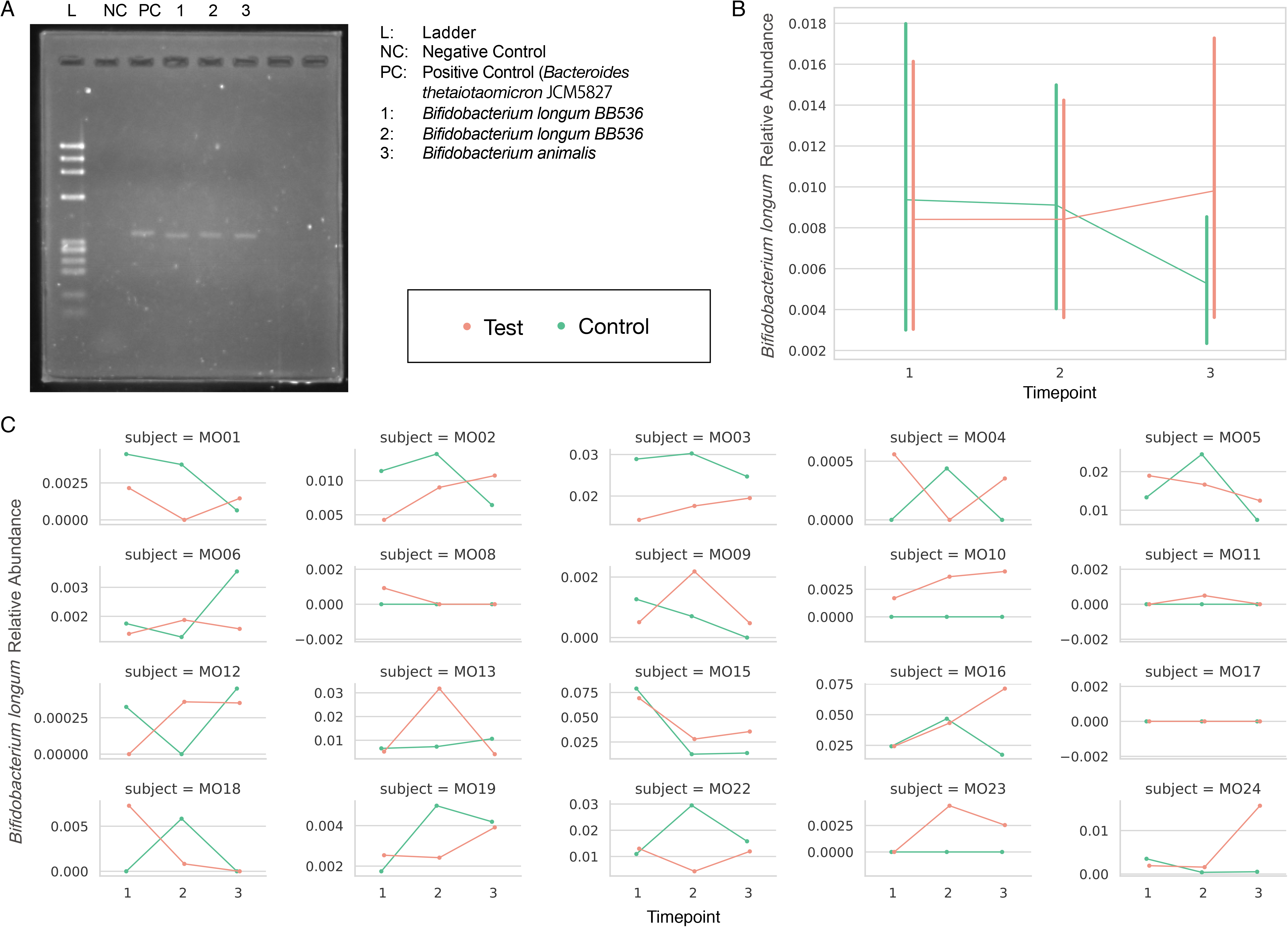

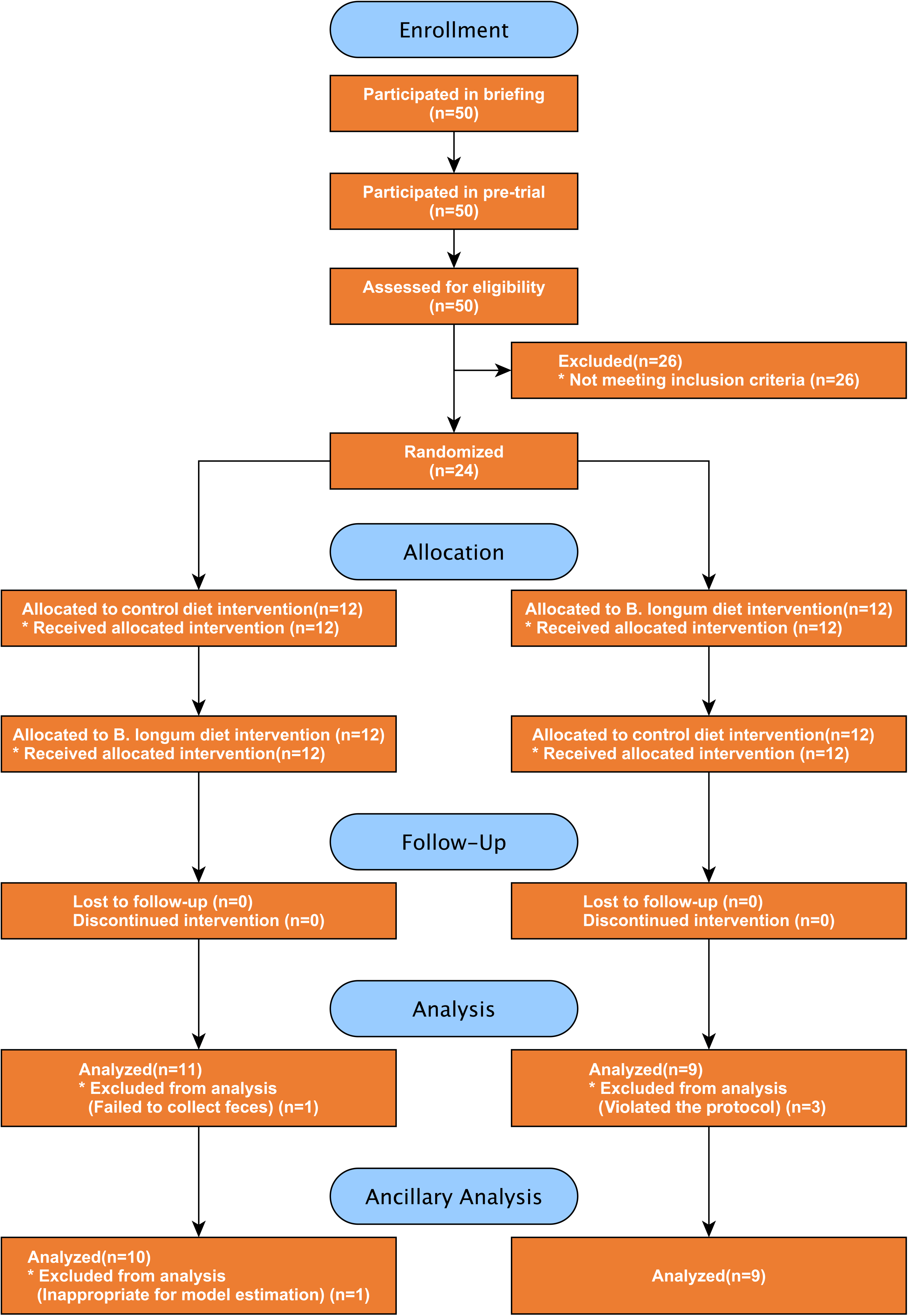

## Notes

### Competing Interest Statement

The authors have declared no competing interest.

### Clinical Trial

UMIN000018924

### Clinical Protocols

https://upload.umin.ac.jp/cgi-open-bin/ctr_e/ctr_view.cgi?recptno=R000021894

